# HPV self-sampling among long-term non-attenders to cervical cancer screening in Norway: A pragmatic randomized controlled trial

**DOI:** 10.1101/2022.06.28.22276756

**Authors:** Gunvor Aasbø, Ameli Tropè, Mari Nygård, Irene Kraus Christiansen, Ingrid Baasland, Grete Alrek Iversen, Ane Cecilie Munk, Marit Halonen Christiansen, Gro Kummeneje Presthus, Karina Undem, Tone Bjørge, Philip E. Castle, Bo Terning Hansen

**Author notes:** **Corresponding author:** Gunvor Aasbø, PhD, Department of Research, Cancer Registry of Norway, Oslo, Norway. Address: Postboks 5313 Majorstuen, 0304 Oslo, Norway.

## Abstract

**Background:** The main aim of the present study is to assess whether human papillomavirus (HPV) self-sampling may increase cervical cancer screening participation among long-term non-attenders in Norway.

**Methods:** A pragmatic randomized controlled trial was initiated in the national cervical screening programme in March 2019. A random sample of 6000 women aged 35-69 years who had not attended screening for at least 10 years were randomized 1:1:1 to receive either (i) a reminder to attend regular screening (control arm), (ii) an offer to order a self-sampling kit (opt-in arm), or (iii) a self-sampling kit unsolicited (opt-out arm).

**Results:** Total screening participation during 6 months following study invitation was 4.8%, 17.0% and 27.7% among women who received a standard reminder letter (controls), women who could order a self-sample kit (opt-in) and women who received a self-sample kit unsolicited (opt-out), respectively (P<0.0001). High-risk HPV was detected in 11.5% of the self-samples and in 9.2% of the clinician-collected samples (P = 0.40). Most women (92.5%) who returned a positive self-sample attended triage. Of 933 women screened in the study (by a clinician or HPV self-sampling), 33 (3.5%) had CIN2+, 31 (3.3%) had CIN3+, and 11 (1.2%) had cervical cancer.

**Conclusion:** We conclude that opt-in and opt-out self-sampling increased screening participation among long-term non-attenders.

**Clinical Trial Registration:** ClinicalTrials.gov NCT03873376

## Background

Screening has reduced cervical cancer incidence (1) and mortality (2) substantially in Norway. However, coverage of the national cervical cancer screening programme has stagnated at a suboptimal 70% (3), and cervical cancer incidence has increased by 14% from the period 2009-2013 to 2014-2018 (3). Under- or unscreened women have an increased risk of cervical cancer (4) and being diagnosed at an advanced stage (5). Thus, interventions that improve screening participation would benefit women’s health.

Approximately 17% of women aged 35-69 years in Norway have not been screened for at least 10 years (3). In Sweden, the detection of high-grade cervical abnormalities among women who had not been screened during the last 10 years was considerably higher than in the general screening population (6), which highlights the need for improved interventions among long-term non-attenders. Important reasons for non-attendance include procrastination, embarrassment, fear of pain and previous negative experiences with the gynaecological exam or a history of sexual abuse (6, 7), as well as practical barriers experienced in everyday life (8). Non-attendance among Norwegian women is also associated with having a male/foreign/young general practitioner (GP) (9), lacking awareness of the recommended screening interval (10), and lower socioeconomic or migrant background (11). Self-sampling for human papillomavirus (HPV) testing, which the women can perform themselves at home, may reinforce the importance of screening, empower women, and mitigate some of the barriers associated with attending a gynaecological screening exam.

HPV infection is a necessary cause of cervical cancer (12). A persistent infection with high-risk HPV (hrHPV) may, through intermediate precancerous stages, lead to cancer (13). Moreover, when cervical cancer is detected at a late stage, effective treatment is limited and prognosis poor (14). When used with HPV assays based on polymerase chain reaction (PCR), self-sampling and HPV testing identifies women with cervical precancer and cancer, cervical intraepithelial neoplasia grade 2 or more severe diagnoses (CIN2+), with similar accuracy as clinician-collected samples (15). Studies also show that self-sampling has a high acceptability among women and is preferred over clinician sampling (16). Opt-out strategies, where women receive a self-sampling kit unsolicited, proved to increase screening participation of under-screened women in a relatively recent meta-analysis (15), and in Norway (17). Opt-in strategies, where women must request a self-sampling kit, have generally not been found to be more effective than invitation letters for increasing screening participation. The effect of self-sampling on participation varies between studies, especially for opt-in strategies, and warrant more studies (15).

Studies on the effect of HPV self-sampling on participation among under-screened women often include women only slightly overdue for screening, who may be easier to engage than long-term non-attenders, while few randomized controlled trials have by design specifically targeted long-term non-attenders (6, 18-21). It is important to gain more knowledge on how secondary cancer prevention among long-term non-attenders may be improved because they may be at a relatively high risk for CIN2+.

### Aim

The primary aim of this study was to compare cervical cancer screening participation among women in Norway who have not attended screening for at least 10 years, who received either a standard reminder letter to attend screening in a clinic (control arm), an offer to order an HPV self-sampling kit (opt-in arm), or were sent an unsolicited HPV self-sampling kit (opt-out arm). As secondary endpoints, we estimated hrHPV positivity rates, attendance to triage after an HPV positive self-sample, and occurrence of CIN2+ among the long-term non-attenders who participated in screening.

## Materials and Methods

### Cervical screening in Norway

The Cancer Registry of Norway (CRN) is responsible for the national cervical cancer screening programme (NCCSP), and invites women aged 25 to 69 years to attend screening. The programme is currently transitioning from cytology every three years to HPV primary screening every five years for women aged 34-69 years. NCCSP uses a centralised invitation procedure and issues standard open reminder letters to women who have not been registered with a screening test during the recommended interval. A second reminder is issued after one year if a woman is still not registered with a test result. These reminders encourage women to schedule an appointment with their GP for a screening test. All cytology and HPV screening results, as well as any associated histology results, are registered in the NCCSP database. All records are associated with the personal identity number unique to each Norwegian resident.

### Study population and design

The target population was women residing in the counties of Hordaland, Rogaland, Sør-Trøndelag and Vest-Agder who had not participated in the screening programme for the last 10 years. These regions consist of mixed rural and urban areas and cover about one-third of the Norwegian population. During the study, primary screening was offered by HPV-test in Hordaland, Rogaland and Sør-Trøndelag, and by cytology in Vest-Agder. A priori power calculations showed that we needed at least 1417 women in each intervention arm to detect a difference of 5 percentage points between the control arm and the intervention arms with 90% power. Using the NCCSP database, we identified 28,125 women who were eligible for the study, of which 6000 (21.3%) were randomly selected for invitation to the study. The randomly selected women were individually randomized 1:1:1 into the control, opt-in or opt-out intervention arm without further restrictions. The eligible women were unaware of the randomization. Blinding was not possible due to the nature of the interventions. Randomization procedures were performed using the sample function in Stata by a programmer who was not involved in the conduct of the trial. Invitations were sent during March-August 2019.

A total of 333 (5.6%) invited women, similarly distributed by intervention arm; 108 (5.4%) in the control arm, 103 (5.1%) in opt-in arm, and 122 (6.1%) in opt-out arm; P = 0.4), were excluded from the study due to: (i) incorrect address, (ii) active refusal to participate in the study, or (iii) ineligibility for the study (living abroad/had prior hysterectomy/had a screening test between study sampling and invitation) (see Figure 1). The trial was registered at ClinicalTrials.gov on 8 March 2019 (NCT03873376).

**Fig. 1.**
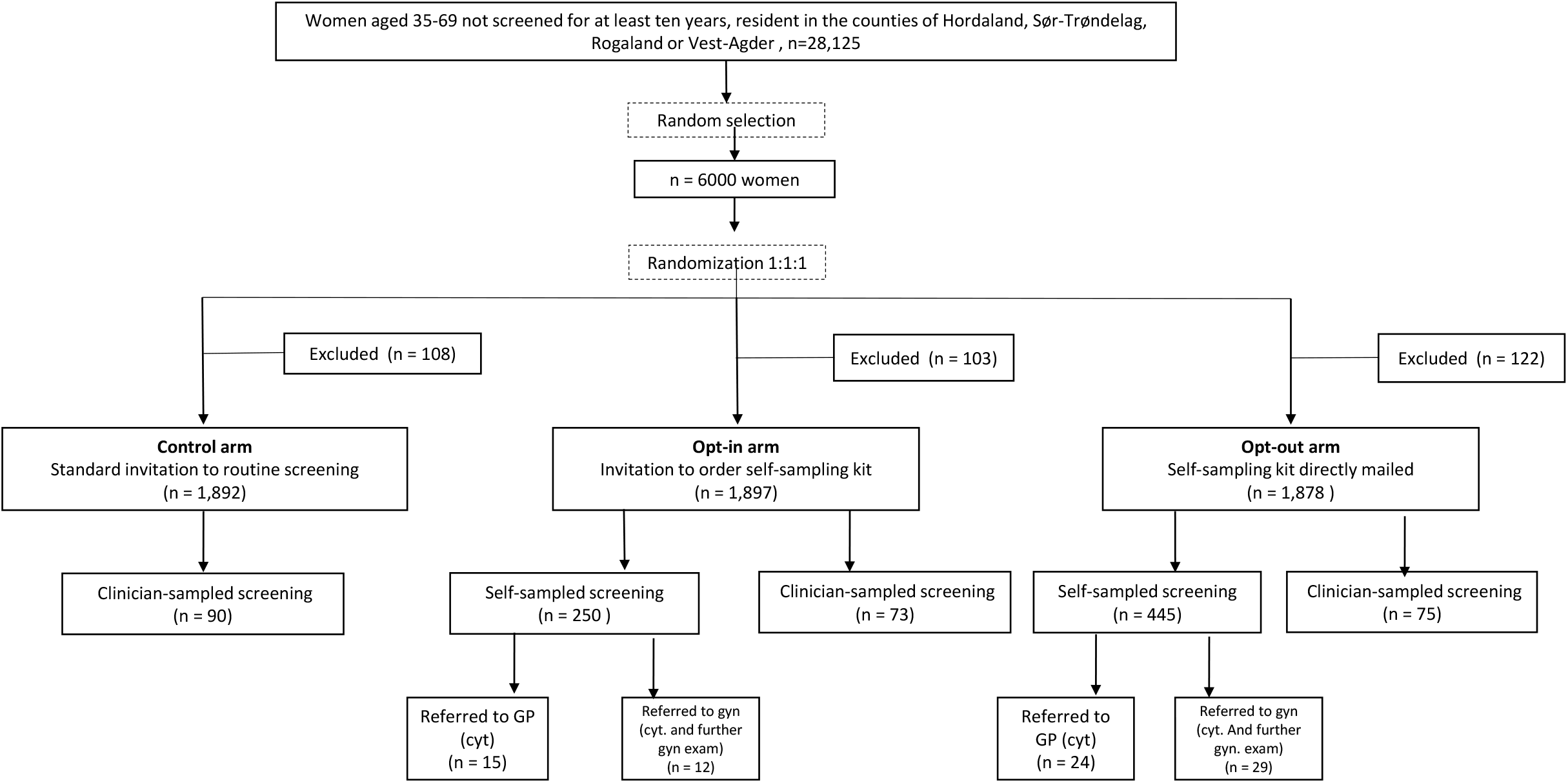
Study flowchart. Excluded women were either not reached, not eligible or declined to participate in the study.

### Invitation letters and information

Women in the control arm received a NCCSP reminder letter to attend regular screening (i.e., encouraging the women to schedule an appointment with their GP). The letter also stated that they were randomly selected to participate in a study aiming to improve cervical cancer prevention. The letter included no information about self-sampling.

Women in the opt-in arm received an invitation letter to order a self-sampling kit. The women could order by mail, e-mail, or through a webpage. The invitation included a unique order code, information about the study, a reply slip, and a pre-paid envelope to return the order. A dedicated webpage for ordering a self-sampling kit was available in Norwegian, English, French, Spanish, Polish, Turkish and Arabic. Each order code could only generate one web order.

A self-sampling kit was mailed unsolicited to women in the opt-out arm, together with the study invitation letter. The content in the opt-in and opt-out invitation letters was the same (apart from the description of the ordering procedure in the opt-in letter) and included information that the women alternatively could schedule a regular screening appointment with their GP if they did not want to use the self-sample.

Similar to regular NCCSP reminders, all invitation letters contained general screening information. The letters were on NCCSP stationary and were signed by the NCCSP head. In addition, letters contained study-specific information, a link to the study webpage, and information that they could refrain from participating in the study. All invitation letters were in Norwegian, except for an English reference to a webpage. The study webpage was available in Norwegian and English and included a link to an instructional video on how to collect a self-sample with options for Norwegian or English audio, and English, French, Spanish, Polish, Turkish or Arabic subtitles.

### The HPV self-sampling kit

The kit included the self-sampling device with user instructions, a sealable plastic bag, a prepaid return envelope addressed to the laboratory, and an information leaflet. The Evalyn® Brush (dry brush) (Rovers Medical Devices B.V, Oss, Netherlands) was used for self-sampling in this study. The self-sampling device had a hidden radio frequency identification chip containing a pseudonymized number unique to each invited woman and had no visible personal information. Women returned the self-sample in the plastic bag and mailed it in the return envelope. Women who had not ordered/returned a self-sample within three weeks received one reminder letter to order/return the self-sample. The self-sampling kit was free of charge for the women.

### HPV detection from self-sampled screening test

All self-samples were analysed at the Norwegian HPV reference laboratory at Akershus University Hospital. Soon after receipt at the laboratory, the brush tip was removed from the self-sampling device, transferred to a vial with 4.5 mL ThinPrep PreservCyt and stored at room temperature for at least 24 hours. During storage, tubes with the brush tip were vortexed three times for 15 seconds to ensure proper dissolution of the sample material. The samples were tested for hrHPV using the cobas® 4800 HPV test (Roche Molecular Systems, Inc, Branchburg, NJ), which individually reports HPV16 and HPV18, in addition to a pooled result of the 12 hrHPV types 31, 33, 35, 39, 45, 51, 52, 56, 58, 59, 66 and 68. Beta-globin is included as internal quality control in a fourth channel. Samples with an invalid test result at first run were retested, which gave a valid test result in every case. Analysis was completed within 21 days after receipt for all samples.

### Clinical management

All women who returned a self-sample were informed about their test result per ordinary mail within six weeks after receipt of the self-sample in the lab. Women with an HPV negative result were encouraged to continue attending the regular cervical cancer screening programme at the recommended interval. Women with a hrHPV positive self-sample received the result together with a pre-booked triage appointment with a physician. To address feasibility and performance of alternative triage strategies that could be implemented in a screening programme, women were sequentially allocated to triage at their GP or a gynaecologist practicing in the largest city in their county. The notification letter to women in each group was identical. Physicians were informed that the patient had tested positive for HPV after participating in a study offering HPV self-sampling to long-term non-attenders. Women referred to GP triage underwent cytology, while women referred to gynaecologist triage underwent further gynaecological examination, which always included cytology and colposcopy, and biopsy if deemed necessary. To simulate a routine healthcare setting, the women paid the deductible for the GP or the gynaecologist appointment themselves (about 30€ and 60€ respectively), as they would in the NCCSP. Management after the scheduled triage visit of women who had a positive hrHPV self-sample was not specific for the study and thus follow national guidelines (22). Similarly, women who had a clinician-collected screening sample (irrespective of treatment arm), or who had a triage cytology with another physician than she was allocated to in the study were managed outside the study. According to the national screening algorithm, women who are HPV16/18 positive are referred to colposcopy with biopsy if the cytology is low- or high-grade, or to a new HPV-test in 12 months if the cytology is normal. Women who are positive for other hrHPV-types are referred to colposcopy with biopsy if cytology high-grade, to a new HPV-test in 12 months if cytology low-grade, or to a new HPV-test in 24 months if cytology normal. Cytology is reported according to the Bethesda system.

### Registry data collection

Data regarding clinician-collected screening tests (including triage tests taken outside the study) was taken from the NCCSP database. Data on histological outcomes from colposcopy referrals were taken from the NCCSP and the CRN databases. Dates were delivered by month and year. If several histological diagnoses were available for the same women, we only considered the most severe diagnosis. The last registry linkage was performed in December 2020, which allowed at least 16 months of registry follow-up after study invitation. Histology diagnoses were interpreted according to WHO guidelines (23). Income data was collected by linkage to Statistics Norway.

### Statistical analysis

Screening participation was defined as returning a valid self-sample or having a clinician-collected screening test within six months after receipt of the invitation letter. Attendance to triage was assessed among women with a hrHPV positive self-sample and was defined as attending at the allocated physician (i.e., GP or study gynaecologist) or an unallocated physician within six months of notification of a positive self-sample. Women who returned a hrHPV positive self-sample and who were not registered with a cytology test by three months following the scheduled appointment, received a reminder from the NCCSP encouraging them to order an appointment at their GP.

We present numbers and proportions of screening participation by intervention arm (control, opt-in, opt-out) overall, by age groups (36-45, 46-55, 56-65, 66-69) and by screening history (time since last screening test 10-15 years, 16-28 years, never). For each self-sampling arm, we present total participation as well as clinician-sampled and self-sampled participation separately. Controls were only screened by a clinician. Differences in screening participation among intervention arms are presented for total participation (i.e., intention to treat), as absolute participation differences (APD, percentage points) and as relative participation differences (RPD, relative risks) with associated 95% confidence intervals (95% CI). Further, we present numbers and proportions of hrHPV positive screening tests and histologically verified high-grade lesions by intervention arm. To investigate the diagnostic yield by mode of triage, we also present the histologically verified high grade lesions separately for women triaged by study gynaecologists, women triaged by study GPs, and women who had a clinician-collected screening test or attended triage outside the study (i.e., women not managed by a study gynaecologist/GP). Since women who have a clinician-collected screening test do not need a separate triage visit, attendance to triage is only presented for women who tested positive by self-sampling. We present attendance to triage by self-sampling arm and by mode of triage (i.e., at gynaecologist or GP). For comparisons of counts and proportions, P-values refer to chi-squared tests, or the Fisher’s exact test for observed counts <5. For comparisons of central tendency, P-values refer to the Mann-Whitney U-test. All tests were two-sided and P-values <0.05 were considered statistically significant. Analyses were performed using Stata version 17MP or R version 3.5.3.

## Results

All baseline characteristics addressed in the study cohort were similarly distributed by study arm (Table 1). The mean age was 54.5, 54.4 and 54.1 years among women in the control, opt-in and opt-out arm. Similar frequency distributions in each study arm were observed for categories of age (P=0.94), time since last screen (P=0.99), county of residence (P=0.96) and income (P=0.28) (Table 1).

**Table 1.** Baseline characteristics of the women in the study population

| Characteristic | Total, all arms<br>*n* (%) |  | Control group<br>*n* (%) |  | Opt-in<br>*n* (%) |  | Opt-out<br>*n* (%) |  |
| --- | --- | --- | --- | --- | --- | --- | --- | --- |
| Total invited | 5,667 | (100) | 1,892 | (100) | 1,897 | (100) | 1,878 | (100) |
| Age (years) |  |  |  |  |  |  |  |  |
| 36 to 45 | 1,318 | (23.3) | 435 | (23.0) | 433 | (22.8) | 450 | (24.0) |
| 46 to 55 | 1,520 | (26.8) | 497 | (26.3) | 520 | (27.4) | 504 | (26.8) |
| 56 to 65 | 1,961 | (34.6) | 663 | (35.0) | 653 | (34.4) | 646 | (34.4) |
| 66 to 69 | 868 | (15.3) | 298 | (15.7) | 292 | (15.4) | 278 | (14.8) |
| Mean age (standard deviation) | 54.3 | (9.9) | 54.5 | (10.0) | 54.4 | (9.8) | 54.1 | (9.9) |
| Time since last screening test |  |  |  |  |  |  |  |  |
| 10 - 15 years | 1,786 | (31.5) | 599 | (31.7) | 590 | (31.1) | 599 | (31.9) |
| Over 15 years | 1,950 | (34.4) | 650 | (34.3) | 658 | (34.7) | 642 | (34.2) |
| Never screened | 1,931 | (34.1) | 644 | (34.0) | 650 | (34.2) | 637 | (33.9) |
| County |  |  |  |  |  |  |  |  |
| Hordaland | 2,106 | (37.1) | 707 | (37.3) | 710 | (37.4) | 689 | (36.7) |
| Rogaland | 1,705 | (30.1) | 568 | (30.0) | 563 | (29.7) | 574 | (30.6) |
| Trøndelag | 1,217 | (21.5) | 415 | (21.9) | 405 | (21.3) | 399 | (21.2) |
| Vest-Agder | 639 | (11.3) | 203 | (10.7) | 220 | (11.6) | 216 | (11.5) |
| Total income (tertiles)<sup>1</sup> |  |  |  |  |  |  |  |  |
| Low | 1,877 | (33.3) | 622 | (33.0) | 632 | (33.5) | 623 | (33.4) |
| Medium | 1,878 | (33.3) | 610 | (32.4) | 659 | (34.9) | 609 | (32.7) |
| High | 1,877 | (33.3) | 650 | (34.5) | 595 | (31.5) | 632 | (33.9) |

<sup>1</sup>Total annual income (wage, welfare payment and capital income) from the most recent Statistics Norway registration during 2014-2018, by tertiles of the total study population. Data missing for 37 women

Total participation (i.e., self-sampled and clinician-sampled tests for the self-sampling arms) was 4.8% in the control arm, 17.0% in opt-in arm and 27.7% in opt-out arm (P < 0.0001, Figure 2, Table 2). Thus, absolute participation differences and relative participation differences in total participation was: 12.3% (95% CI 10.3 - 14.2) and 3.6 (95% CI 2.9 - 4.5), respectively for opt-in vs. controls; 22.9% (95% CI 20.7 - 25.2) and 5.8 (95% CI 4.7 - 7.2), respectively, for opt-out vs. controls; and 10.7% (95% CI 8.0 - 13.3) and 1.6 (95% CI 1.4 - 1.8), respectively, for opt-out vs. opt-in. Differences in participation between intervention arms were largely due to self-sampling use, since there was similar attendance to clinician-collected screening: 4.8% for the control arm, 3.8% for opt-in, and 4.0% for opt-out (P = 0.33, Table 2).

**Fig. 2.**
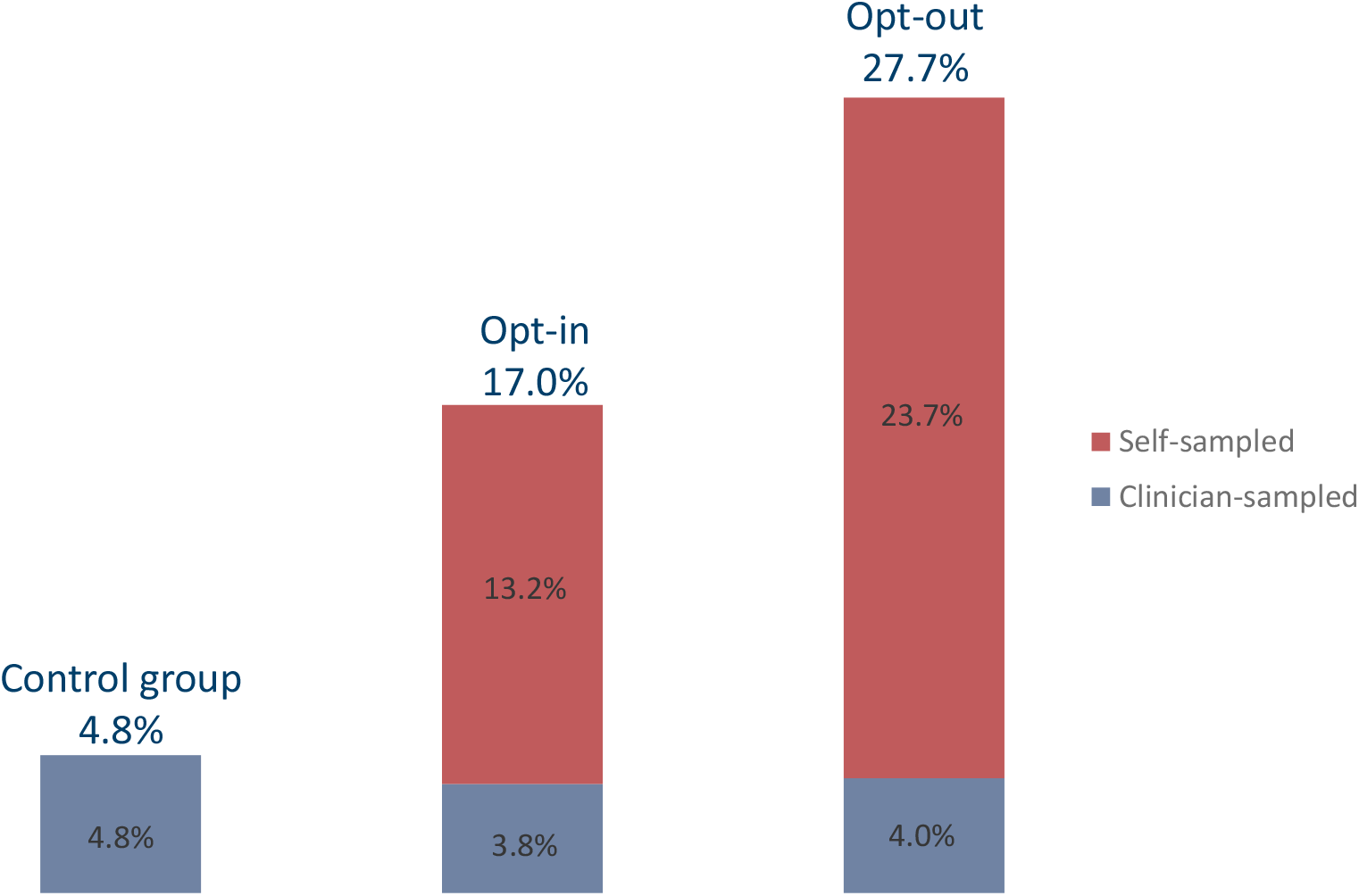
Participation rate (%) during six months following the invitation.

**Table 2.**
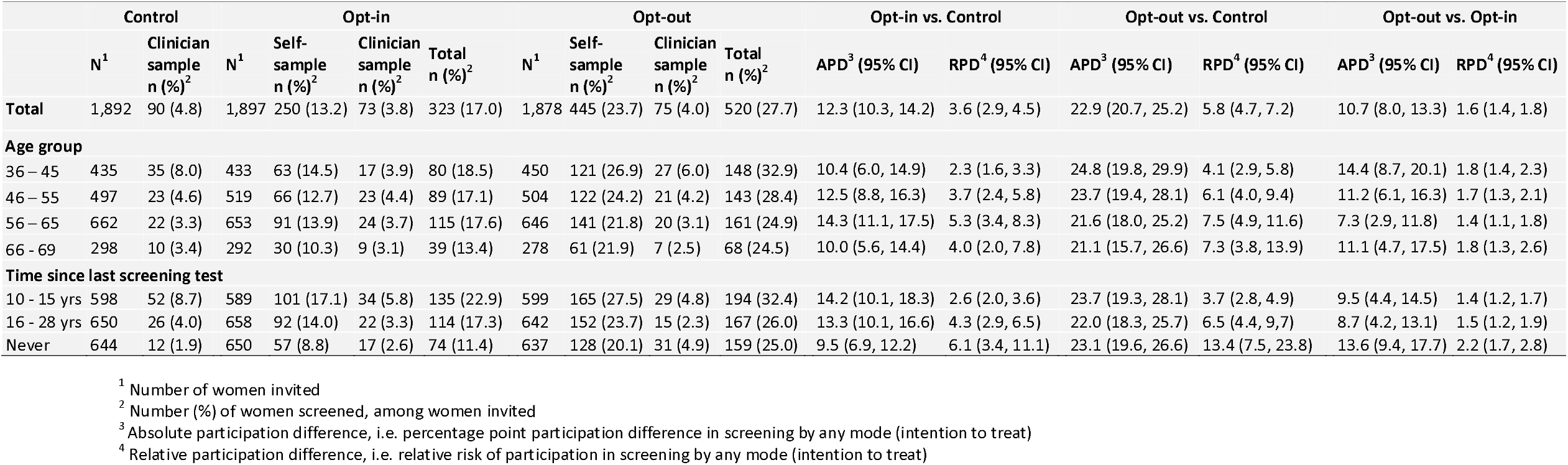
Screening participation among long-term non-attenders by intervention arm, and contrasts in any participation

In the opt-in arm, 403 out of 1897 (21.2%) women ordered a self-sample, of which 144 (35.7%) ordered after receiving a reminder, and 250 (13.2%) returned the self-sample for analysis. Opt-in orders were made by ordinary mail (51.6%), web (33.0%), e-mail (14.2%) or telephone to the study centre (1.2%; not presented as an option in the invitation letter). Among 445 women in the opt-out arm who submitted a self-sample, 254 (57.1%) did so after receiving a reminder.

Self-sampling increased total participation in all age groups and screening history categories. Total participation was consistently highest in the opt-out arm, intermediate in the opt-in arm and lowest in the control arm (Table 2). In each arm, total participation was highest in the youngest age group (age 36-45 years), at 8.0%, 18.5% and 32.9% among controls, opt-in and opt-out, respectively, and tended to decrease with age (P-trend in total participation by age group: 0.001, 0.15 and 0.002 for controls, opt-in and opt-out, respectively). Participation generally decreased with increasing time since last screening test, and this pattern was evident for both clinician-sampling and self-sampling, and in each intervention arm (P-trend in total participation by time since last screening test (10-15 years, 16-28 years or never screened): <0.0001, <0.0001 and 0.004 for controls, opt-in and opt-out, respectively). Among women who never had been screened previously, total participation was 1.9% for controls, 11.4% for opt-in and 25.0% for opt-out (P < 0.0001). Also, the RPD was highest among never screeners, at 6.1 (95% CI 3.4 - 11.1), 13.4 (95% CI 7.5 - 23.8) and 2.2 (95% CI 1.7 - 2.8) for opt-in vs. controls, opt-out vs. controls and opt-out vs. opt-in, respectively (Table 2).

Among all women who were tested for HPV, 11.0% were positive for any hrHPV. The hrHPV positivity rate was slightly higher for self-sampled tests than for clinician-collected tests, at 11.5% and 9.2% respectively (P = 0.40, Table 3). Among controls, 6.0% were positive for any hrHPV, which was non-significantly lower than observed among women in the opt-in (10.8%) and opt-out (11.8%) intervention arms (P = 0.28) (Table 3).

**Table 3.** Proportion of hrHPV at screening^1^ among long-term non-attenders in total and by intervention arm

| Type of HPV screening test | All women screened by HPV-test |  |  |  | Control |  |  |  | Opt-in |  |  |  | Opt-out |  |  |  |
| --- | --- | --- | --- | --- | --- | --- | --- | --- | --- | --- | --- | --- | --- | --- | --- | --- |
|  | N | Any hr positive<sup>2</sup> n(%) | 16/18 positive<sup>3</sup> n(%) | Other hr positive<sup>4</sup> n(%) | N | Any hr positive<sup>2</sup> n(%) | 16/18 positive<sup>3</sup> n(%) | Other hr positive<sup>4</sup> n(%) | N | Any hr positive<sup>2</sup> n(%) | 16/18 positive<sup>3</sup> n(%) | Other hr positive<sup>4</sup> n(%) | N | Any hr positive<sup>2</sup> n(%) | 16/18 positive<sup>3</sup> n(%) | Other hr positive<sup>4</sup> n(%) |
| Total | 913 | 100(11.0) | 38(4.2) | 60(6.6) | 84 | 5(6.0) | 0(0.0) | 4(4.8) | 314 | 34(10.8) | 11(3.5) | 23(7.3) | 515 | 61(11.8) | 27(5.3) | 33(6.4) |
| Clin-sample | 218 | 20(9.2) | 5(2.3) | 13(6.0) | 84 | 5(6.0) | 0(0.0) | 4(4.8) | 64 | 7(10.9) | 2(3.1) | 5(7.8) | 70 | 8(11.4) | 3(4.3) | 4(5.7) |
| Self-sample | 695 | 80(11.5) | 33(4.8) | 47(6.8) | - | - | - | - | 250 | 27(10.8) | 9(3.6) | 18(7.2) | 445 | 53(11.9) | 24(5.4) | 29(6.5) |

<sup>1</sup> Women who attended clinician screening with cytology only (n=20) are not included. Six of these were in the control group, nine in the opt-in group and five in the opt-out group.
<sup>2</sup> Positive for one or more of the hrHPV-types 16, 18, 31, 33, 35, 39, 45, 51, 52, 56, 58, 59, 66 or 68. Two women (one control and 1 opt-out) were registered as any hrHPV positive, but lacked information on hrHPV-type
<sup>3</sup> Positive for HPV16 and/or 18, and may be positive for other hrHPV-types
<sup>4</sup> Positive for one or more of the hrHPV-types 31, 33, 35, 39, 45, 51, 52, 56, 58, 59, 66 or 68, but negative for 16/18

Triage attendance was similar in the opt-in and opt-out arms, at 77.8% and 79.3% (P = 1), respectively, for scheduled attendance, and 92.6% and 92.5% (P = 1), respectively, for any attendance within six months (Table 4). Attendance for women allocated to gynaecologist triage (irrespective of intervention arm) for scheduled and any attendance was 75.6% and 90.2%, respectively, which was non-significantly lower than observed among women allocated to GP triage, who had corresponding attendance of 82.1% and 94.9% (P = 0.67 for scheduled attendance, P = 0.72 for any attendance, Table 4). The median (interquartile range (IQR)) period between a positive self-sample and the scheduled triage appointment was shorter for women allocated to GP triage than gynaecologist triage, at 19 (IQR 14, 25) and 43 (IQR 29, 54) days (P < 0.0001), respectively. However, among women who were biopsied and had a histology result, the median (IQR) period between the positive self-sample and the histological diagnosis was longer for women who were allocated to and attended GP triage than for women who were allocated to and attended gynaecologist triage, at 4 (IQR 3, 8) and 3 (IQR 2, 3) months, respectively (P = 0.01).

**Table 4.** Attendance to triage among long-term non-attenders with a hrHPV positive self-sample^1^

|  | Total, n (%) | Opt-in, n (%) | Opt-out, n (%) |
| --- | --- | --- | --- |
| **Total** |  |  |  |
| Positive tests | 80 (100.0) | 27 (100.0) | 53 (100.0) |
| Had triage test<sup>2</sup> | 63 (78.8) | 21 (77.8) | 42 (79.3) |
| Had triage test<sup>3</sup> | 74 (92.5) | 25 (92.6) | 49 (92.5) |
| **Allocated gynaecologist** |  |  |  |
| Positive tests | 41 (100.0) | 12 (100.0) | 29 (100.0) |
| Had triage test<sup>2</sup> | 31 (75.6) | 10 (83.3) | 21 (72.4) |
| Had triage test<sup>3</sup> | 37 (90.2) | 11 (91.7) | 26 (89.7) |
| **Allocated RGP** |  |  |  |
| Positive tests | 39 (100.0) | 15 (100.0) | 24 (100.0) |
| Had triage test<sup>2</sup> | 32 (82.1) | 11 (73.3) | 21 (87.5) |
| Had triage test<sup>3</sup> | 37 (94.9) | 14 (93.3) | 23 (95.8) |

<sup>1</sup>Positive clinician-samples do not require attendance for triage and are hence not included in this table
<sup>2</sup> Attended follow-up within 6 months of notification of positive HPV-result, at allocated physician
<sup>3</sup> Attended follow-up within 6 months of notification of positive HPV-result, regardless of where the exam took place

A total of 1, 12, and 20 prevalent cases of CIN2+ were detected in the control, opt-in, and opt-out arms, respectively (Table 5). Thus, we detected CIN2+ in 0.1%, 0.6% and 1.1% of the invited women (P = 0.0002), and in 1.1%, 3.7% and 3.8% of the screened women (P = 0.52) in the control, opt-in, and opt-out arms, respectively. A total of 0, 4 and 7 cases of cervical cancer were detected in the control, opt-in and opt-out arms, respectively, giving corresponding detection rates per woman screened of 0%, 1.2% and 1.3% (P = 0.81). In each of the self-sampling arms, the detection rate of CIN2+ and cancer per woman screened was similar among women who screened by self-sampling and women who attended screening at a clinic (Table 5).

**Table 5.** Histologically verified high-grade lesions^1^ among long-term non-attenders in total and by intervention arm

|  | Total, n (%) | Control, n(%) |  | Opt-in, n(%) |  |  | Opt-out, n(%) |  |
| --- | --- | --- | --- | --- | --- | --- | --- | --- |
|  |  | Clinician-sample | Total | Self-sample | Clin-sample | Total | Self-sample | Clin-sample |
| Invited | 5,667 | 1,892 | 1,897 |  |  | 1,878 |  |  |
| Screened | 933 (16.4) | 90 (4.8) | 323 (17.0) | 250 (13.2) | 73 (3.8) | 520 (27.7) | 445 (23.7) | 75 (4.0) |
| Had histology result<sup>2</sup> | 61 (1.1) | 3 (0.2) | 20 (1.1) | 15 (0.8) | 5 (0.3) | 38 (2.0) | 32 (1.7) | 6 (0.3) |
| Had histology result<sup>3</sup> | 61 (6.5) | 3 (3.3) | 20 (8.6) | 15 (6.0) | 5 (6.8) | 38 (7.3) | 32 (7.2) | 6 (8.0) |
| CIN2+ detected<sup>2</sup> | 33 (0.6) | 1 (0.1) | 12 (0.6) | 9 (0.5) | 3 (0.2) | 20 (1.1) | 17 (0.9) | 3 (0.2) |
| CIN2+ detected<sup>3</sup> | 33 (3.5) | 1 (1.1) | 12 (3.7) | 9 (3.6) | 3 (4.1) | 20 (3.8) | 17 (3.8) | 3 (4.0) |
| CIN3+ detected<sup>2</sup> | 31 (0.5) | 1 (0.1) | 10 (0.5) | 7 (0.4) | 3 (0.2) | 20 (1.1) | 17 (0.9) | 3 (0.2) |
| CIN3+ detected<sup>3</sup> | 31 (3.3) | 1 (1.1) | 10 (3.1) | 7 (2.8) | 3 (4.1) | 20 (3.8) | 17 (3.8) | 3 (4.0) |
| Cervical cancer detected<sup>3</sup> | 11 (1.2) | 0 (0.0) | 4 (1.2) | 3 (1.2) | 1 (1.4) | 7 (1.3) | 6 (1.3) | 1 (1.3) |

<sup>1</sup> CIN2, CIN3, ACIS or cancer
<sup>2</sup> Per cent of women invited
<sup>3</sup> Per cent of women screened

A relatively high proportion of women with screening-positive tests were subsequently diagnosed with CIN2+ (Table 6). Although the percentage of histologic diagnoses was higher in the group attending gynaecologist triage, the CIN2+ yield was similar in this group when compared to women attending GP triage and women who were screened in a clinic or were triaged outside the study, at 32.3%, 34.4% and 35.3% of the screening-positive tests, respectively. Similar occurrence was observed for CIN3+, thus few of the histologically verified cases were CIN2, an equivocal diagnosis of precancer (24). Cancer was diagnosed in four women who attended gynaecologist triage, four women who attended GP triage, and three women who attended screening at a clinic or triage outside the study, which constituted 12.9%, 12.5% and 8.8% of the screening-positive tests respectively. For all screening and triage modes, the CIN2+ yield was considerably higher among women who tested positive for HPV16/18 than among women who were positive for other hrHPV types. Among women who screened positive for HPV16/18, the percentage of CIN2+ was 64.3%, 55.6% and 69.2% for women attending gynaecologist triage, GP triage, and women who attended screening at a clinic or triage outside the study, respectively (Table 6).

**Table 6.**
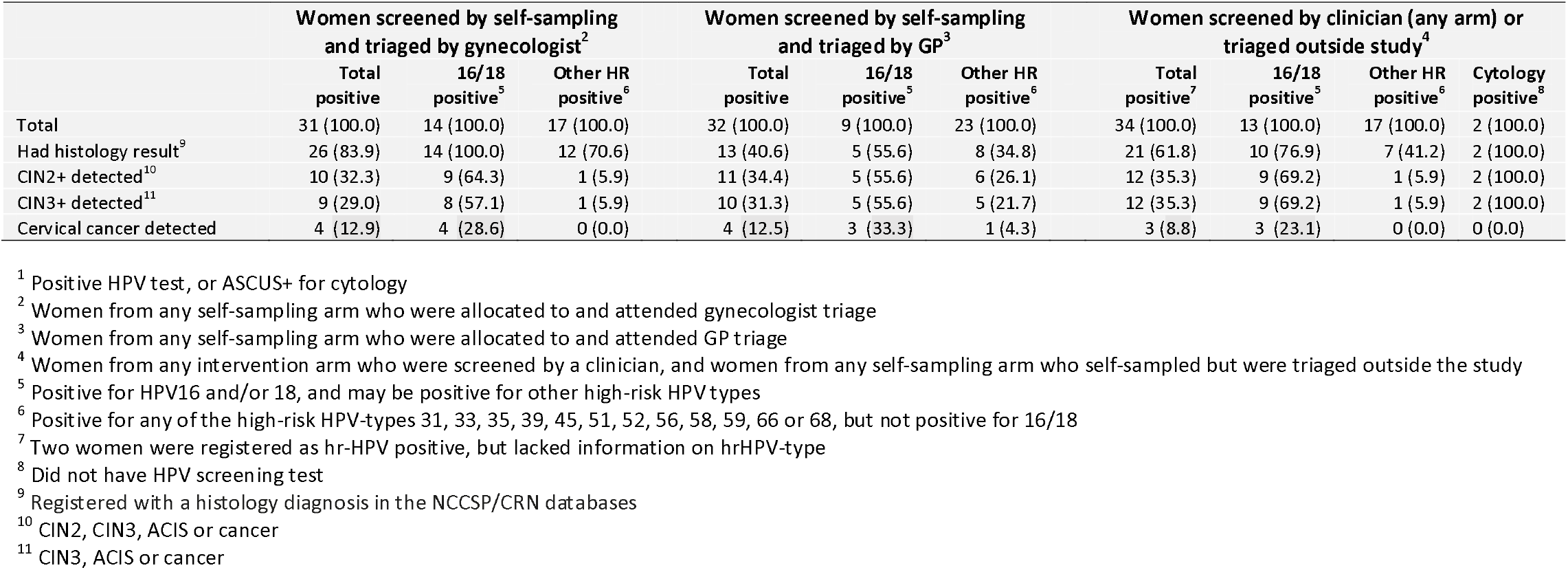
Histologically verified high-grade lesions (n, % of total) among screening test positive^1^ women by triage mode

## Discussion

We show that offering HPV self-sampling to long-term non-attending women as opt-in or as opt-out significantly increases the cervical screening participation relative to the standard reminder letter, and that opt-out gives the largest increase in participation.

Although the effect of self-sampling interventions may differ between studies due to differences in study design and screening setting, increased participation by the opt-out self-sampling strategy is well established compared to clinician-collected screening (15, 25). In the present study, we observed larger absolute and relative differences in participation between the opt-out self-sampling arm and the controls than was reported in a recent meta-analysis of self-sampling (15). Moreover, we observed significantly increased participation for the opt-in strategy, which was not found in the meta-analysis (15). Our results regarding self-sampling participation generally coincide with randomized controlled trials that have targeted long-term non-attenders (6, 18-20) or show stratified analyses for this part of the screening population (21, 26).

Characteristics of the targeted screening population are important for the way in which self-sampling interventions may affect participation. When self-sampling is offered more widely to women who are overdue for screening, it tends to replace screening at a clinic in addition to increase overall participation (17, 21). We observed that the clinician-collected screening uptake was only marginally lower in the self-sampling arms than in the control arm, indicating that self-sampling hardly replaces clinician-collected screening when offered exclusively to long-term non-attenders. Our observation that some long-term non-attending women offered self-sampling still preferred to be screened by a clinician, demonstrates the importance of maintaining this option even if self-sampling is implemented in the Norwegian screening programme. Overall, we report similar effects on participation as a Swedish study targeting long-term non-attenders (6). However, the absolute increases in participation for the self-sampling interventions versus controls were considerably higher in our study, but the relative participation difference was lower, which may relate to the slightly different screening contexts experienced by long-term non-attending women in these countries. For instance, the Swedish screening programme issues far more screening reminders than the Norwegian programme and offers prescheduled screening appointments rather than an encouragement to order an appointment for screening, which in part may explain the higher coverage observed for ordinary screening in Sweden (4) and the very low participation to clinician-collected screening among long-term non-attenders (6). On a general note, groups that respond poorly to the control intervention, i.e. the standard invitation to attend screening in a clinic, have a high potential to benefit from self-sampling in relative terms, as we can observe in the relative participation difference estimates of the never-screeners and the women in the oldest age brackets in our study population.

Although inferior to opt-out, we found that opt-in also increased screening participation when compared to controls who received the routine reminder letter. Thus, this type of intervention should not be dismissed, although opt-in in other settings often did not improve participation beyond a routine reminder letter to screen (15). The way in which this intervention is offered may be of particular importance for achieving increased participation. The present study results suggest that alternative ordering options that include ordinary mail and a web-solution may be considered for opt-in among long-term non-attenders, and that a reminder to order the self-sampling device may be effective. Some effect of opt-in has also been reported in intention-to-treat analyses of other randomized controlled trials (6, 21) and observational studies (27, 28) in Scandinavia.

Long-term non-attending women may be hesitant to undergo a gynaecological examination for screening purposes (29), but our study showed that the vast majority attended to examination by a physician if they submitted a positive self-sample. The attendance to triage observed among women with a positive hrHPV self-sample was somewhat higher than attendance to follow-up after positive screening tests in the NCCSP (3). A high attendance to triage after a positive self-sample has also been found in other settings (15). We found that triage attendance was similar for women who were scheduled for GP and gynaecologist triage. However, some women chose to be triaged by another physician than they were allocated to in the study, and this happened slightly more often among women allocated to a gynaecologist than among women allocated to their GP. One reason for this difference could be that the gynaecologists on average were located further away from the women than their GP. Moreover, we observed a longer time lag between notification and the scheduled appointment for women allocated to a gynaecologist, which could have compelled more of these women to reschedule to a physician who could offer an earlier appointment.

The hrHPV positivity rate of the total study population of long-term non-attenders was nearly twice as high as the corresponding rate among women of the same age in the ordinary screening population (3). In both self-sampling arms, women who chose to attend screening at a clinic and women who chose to use self-sampling had similar hrHPV positivity rates, indicating that the mode of screening chosen by long-term non-attending women is not associated with risk of infection.

Due to low numbers, the CIN2+ occurrence observed here should be interpreted with caution, especially in terms of comparisons between intervention arms or other subgroups. Among all the long-term non-attending women who were screened during the study, 3.5% and 1.2% were diagnosed with CIN2+ and cervical cancer, respectively, while the corresponding rate among women aged 34-69 years in the NCCSP is 1.2% and 0.1% (3). Thus, CIN2+ and cervical cancer yield among long-term non-attenders observed in this study was very high compared to the ordinary screening population, which probably reflects a real difference in risk. However, surveillance bias could also have contributed since most physicians who triaged women who had submitted a hrHPV-positive self-sample knew that they were long-term non-attenders and part of a study, and women who were triaged by a study gynaecologist underwent colposcopy at the triage visit. However, the occurrence of CIN2+ among women with a positive screening test (hrHPV or cytology) was very similar regardless of differences in diagnostic follow-up.

GP and gynaecologist triage of long-term non-attending women who had a hrHPV-positive self-sample gave similar attendance and diagnostic yield. However, for long-term non-attending women who have CIN2+, it is important to be treated quickly, and our results clearly showed that the final diagnostics appeared earlier if women were referred directly to a gynaecologist who could perform a colposcopic biopsy at the triage visit than if they were referred to their GP for cytology triage. Furthermore, the CIN2+ and CIN3+ detection rates among long-term non-attenders with a positive screening test exceeded the risk thresholds for which colposcopy usually is considered good practice (30, 31).

### Strengths and limitations

A strength of the present study is the use of national registry data that ensures a precise definition of the study cohort and complete information on participation and diagnostics. Targeted sampling of long-term non-attenders also ensures a sufficient sample size for robust inference regarding comparisons of participation by study arm in this part of the screening population. Another strength is the randomized design that enhances the representativeness of the study sample and the similarity of the intervention arms. The intervention arms should thus be highly comparable, and the results of the present study should have high generalizability to the population of long-term non-attenders in Norway. Furthermore, we used a self-sampling device and a PCR-based HPV DNA test that were clinically validated and gave no invalid self-samples in this study. Finally, the trial was embedded in the NCCSP, which should make the study results relevant to a real-world scenario where self-sampling is offered as part of an organized programme. However, the invited women were informed that this was a study, which could have influenced their motivation to participate. A further limitation is the relatively low subgroup numbers regarding hrHPV positivity, attendance to triage and CIN2+ occurrence, which limits the precision of the inference that can be made regarding these secondary outcomes of the study.

## Conclusion

We conclude that opt-in and opt-out self-sampling strategies increase screening participation among long-term non-attenders to cervical screening in Norway. If implemented as part of the screening programme, both strategies would probably improve secondary prevention of cervical cancer and thus benefit women’s health. Long-term non-attenders also have an elevated risk for hrHPV infection and high-grade cervical lesions, which highlights the large potential to improve cervical cancer prevention by increasing the screening participation of this population. The opt-out strategy would maximize the preventive effect because it clearly increased participation the most. Maintaining an offer for clinician-collected screening will still be important, since some long-term non-attenders offered self-sampling preferred this option. We also show that management of hrHPV-positive self-samples by GP cytology triage or direct referral to colposcopy by gynaecology specialists give similar results in terms of triage attendance and diagnostic yield. Our study supports findings that direct referral to colposcopy might be the best option for hrHPV-positive women in this population, especially if they are positive for HPV16/18, because they are at a relatively high risk for sequelae and colposcopy referral gives the shortest lag to a histologically confirmed diagnosis and treatment. Colposcopy is generally a safe procedure, thus over-referral will not be a burden for the patient.

## Supporting information

Consort checklist

## Data Availability

The data contains personal information and the study participants have not consented to public data sharing. Data access requires permission by relevant Norwegian authorities.

## Additional Information

## Acknowledgements

We thank Mona Hansen for valuable contribution in the planning and preparation of procedures for sample and data handling at the laboratory; Suzanne Campbell and Kristina S. Gjøtterud for data management; Kristin H. Brenden, Gintaras Pikelis, Håkan Olofsson, Caroline Skudal and Björn Eklund for IT design and support; Randi Waage and Gry B. Skare for NCCSP support; Pia C.M. Osborne and Minh T. Le for data collection assistance, and Jeanette Hoel who was the user representative of the study.

## Authors’ contributions

BTH, MN and AT conceived the study. GA, IKC, TB and PEC contributed to the study design. BTH was the principal investigator. BTH, GA and KU coordinated the execution of the trial. IKC supervised sample flow and HPV-testing. BTH and GA were responsible for data analyses. IB, GAI, ACM, MHC and GKP contributed to data collection. GA wrote the first manuscript draft with support from BTH. All authors commented on drafts and approved the final manuscript.

## Ethics approval and consent to participate

The South-Eastern Committee A for Medical and Health Research Ethics in Norway approved the study (project-ID: 2019/111) and Oslo University Hospital Data Protection Officer approved the processing of data in the study (project-ID: 18/14056). In addition, a statement regarding the processing of personal data was also obtained from the Data Protection Officer at Akershus University Hospital. For women in self-sampling arms, the invitation included information that by returning the self-sample, they consented to the subsequent procedures described in the invitation letter. All invited women, including those in the control arm, were informed that they could withdraw from the study at any time. The study was performed in accordance with the Declaration of Helsinki

## Consent for publication

Not applicable

## Competing Interests

Dr. Castle has received HPV tests and assays for research at a reduced or no cost from Roche, Becton Dickinson, Cepheid, and Arbor Vita Corporation. All other authors declare no conflict of interest.

## Funding information

This study was funded by the Norwegian Cancer Society and Thea Steen Memorial fund (grant number 182687-2016). The funding source had no role in study design, data collection and analysis, preparation of the manuscript, or decision to publish.

## Disclaimer

The opinions expressed by the authors are their own and this material should not be interpreted as representing the official viewpoint of the U.S. Department of Health and Human Services, the National Institutes of Health, or the National Cancer Institute.

## Notes

### Clinical Trial

NCT03873376

